# Returning to the workplace during the COVID-19 pandemic: The concerns of Australian workers

**DOI:** 10.1101/2021.03.28.21254520

**Authors:** Daniel Griffiths, Luke Sheehan, Caryn van Vreden, Peter Whiteford, Alex Collie

## Abstract

**Purpose:** To determine the nature and prevalence of workers’ concerns regarding workplaces reopening during the pandemic. To identify characteristics of workers and industries where particular concerns are more common.

**Methods:** Prospective cohort study of 1063 employed Australian adults, enrolled at the start of the pandemic. Data on attitudes to workplaces reopening were collected 1 July – 30 September 2020. The frequency of concerns describes infection risk and changes to work and impact on home life. Regression models examined associations between demographic and industry factors with reopening concerns.

**Results:** More than four in five (82.4%) of workers reported concerns about workplace infection risk. Just over half (53.4%) reported concerns about impacts to work and home life. Concerns were more prevalent for workers reporting psychological distress, financial stress, and among those exclusively working from home. Concerns regarding infection risk were common for workers in health care (IRR=1.16, 95% CI=[1.01, 1.33]), retail (IRR=1.31, 95% CI=[1.06, 1.61]), and accommodation/food service industries (IRR=1.25, 95% CI=[1.01, 1.55]). Concerns regarding changes to work and home life were more common for female workers (IRR=1.24, 95% CI=[1.07, 1.43]), and partners/spouses with dependent children (IRR=1.44, 95% CI=[1.16, 1.79]).

**Conclusion:** Concerns of COVID-19 infection in the workplace are common. Many workers are also concerned about changes to their work and home life. The prevalence of concerns is related to the nature of work and responsibilities at home. Actions that reduce risk of workplace transmission, coupled with effective communication of infection controls, may alleviate worker concerns whilst recognising workers’ family and social circumstances.

## Introduction

It is clear that workplaces have been a major source of COVID-19 transmission, and that some workplaces are higher risk than others [1]. Strategies to reduce workplace transmission have ranged from the introduction of infection control measures [2] through to business closures, sometimes with government financial support for affected businesses, individuals or both. Where possible business transitioned to remote or virtual working arrangements, more common in high income countries [3]. Accordingly, there has been a global shift in the way work is conducted, with large sections of the workforce spending extended periods of time away from their ‘usual’ or pre-COVID workplace, or working at their usual workplace but under substantially modified working conditions. Following periods of lockdown, businesses and workplaces will begin to reopen. In some nations infection rates are low, enabling a gradual return to workplaces across most industries. Community-wide or workplace-based immunisation programs may also enable more workers to return to the workplace.

Several changes to work and workplaces may leave workers concerned about their future. Some workers may be concerned about the risk of occupational infection. Others may have adjusted to a new way of working and be concerned about losing the benefits of their modified working situation, such as more time spent with family. Still others may be concerned that their job role will have changed or about their ability to work productively under modified arrangements. Understanding the nature of workers’ concerns, and identifying those most concerned, will assist businesses and governments to more effectively address and alleviate workers concerns as the pandemic unfolds.

Infection risk is not equitable amongst workers [4]. Infection risk leading to COVID-19 has been highlighted for workers in essential occupations such as health and aged care workers, bus drivers and meat-processing workers [5, 6], many of whom interact in-person with members of the public or closely other workers and have been required to continue work at their usual workplaces throughout the pandemic period. Individuals working in indoor settings such as offices may be susceptible to airborne transmission, or on public transportation when commuting to and from work [7]. Risk of infection, and the perception of risk, is also influenced by outbreaks of COVID-19 within workers’ localities. For example, in Australia, the setting of this study, infection risk increased during a second wave of COVID-19 localised within the state of Victoria in mid-2020, which led to large-scale business closures and a working from home directive for many industries in that state [8].

The impacts of working from home are well described and include reported improvements in quality of life, more time with family, as well as some potential disadvantages such as work intensification and less ability to switch off [9]. As workplaces reopen, at-home workers may risk losing advantages like increased time with family and friends and fewer transportation costs, while others may desire return to the workplace in order to reduce perceived or actual negative consequences of working from home.

Workplaces are highly diverse settings. Understanding the extent of worker concerns, and characteristics of individuals who are most concerned about workplaces reopening during the COVID-19 can inform evidence-based strategies to help address and alleviate those concerns.

## Methods

### Design, setting and participants

We report findings from a prospective cohort study on work loss and health during the COVID-19 pandemic [10, 11]. Participants were employed in a paid job prior to the COVID-19 pandemic, or were self-employed, and were aged 18 years or older, residing in Australia. Participants enrolled into the study between 27 March and 12 June 2020, and completed a 20-minute baseline survey (either via online or a telephone survey). This study reports findings from the third survey of the study, conducted 3 months after the baseline survey, with data collected between 1 July and 30 September 2020. During this time period workplaces around Australia were gradually reopening following a national lockdown during the March to May 2020 period, with the exception of the state of Victoria which was in the midst of a second wave of COVID-19 cases and an extended community lockdown [12]. For the current analysis, the cohort was restricted to those participants who reported being employed at the third survey and answered questions on workplaces reopening concerns. This sub-cohort encompassed individuals that were either working from home, working partly from home and partly at their usual workplace, working exclusively at their usual workplace, or stood-down from work (i.e. employed but not currently working). Participants who were unemployed at the third survey were not asked these questions, and therefore not included. Additionally, participants missing data for independent or dependent variables used in regression modelling were excluded.

### Concerns regarding workplaces reopening

Participants were presented with the following question *‘Thinking about workplaces reopening, are you worried about any of the following things?’* along with a list of fourteen items (Table 1) each with a binary response option (yes, no). Response items were developed by the study investigator team to reflect issues being raised in public discussions of workplace reopening, and identified in early reports from published academic literature [1].

**Table 1.** Concerns of workers as workplaces reopen.

|  | **Workers with concern N (%)** |
| --- | --- |
| **Overall concern** | 917 (86.3) |
| **Infection risk** | 876 (82.4) |
| Getting infected with COVID-19 | 525 (49.4) |
| Work colleagues coming to work while sick | 521 (49.0) |
| Infecting someone else with COVID-19 | 520 (48.9) |
| Maintaining physical distance from members of the public | 488 (45.9) |
| Maintaining physical distance from my work colleagues | 365 (34.3) |
| Appropriate cleaning of my workplace | 311 (29.3) |
| Travelling to and from work on public transport | 253 (23.8) |
| Access to soap or hand sanitiser | 135 (12.7) |
| **Impact on work and home life** | 568 (53.4) |
| Reduction in the time I can spend with family / friends | 251 (23.6) |
| How changes to the workplace will affect my ability to work | 244 (23.0) |
| Uncertainty around what my job will involve when I go back | 185 (17.4) |
| Impact on my roles and responsibilities at home | 168 (15.8) |
| Pressure to continue working even when feeling sick | 156 (14.7) |
| Losing the ability to work from home | 131 (12.3) |

### Independent variables

Independent variables were extracted from the study database and included data items describing workers’ demographics, residence, work details (prior to the pandemic and current), health, finances, and social interactions, and survey mode. Demographic factors included gender, age group, the highest level of education attained, household composition and state of residence, all collected at the baseline survey. Details of occupation, industry and employment type were also collected at the baseline survey and referred to participants work prior to the pandemic. Response categories with fewer participants were combined to form larger groups with the exception of gender, where included gender responses were binary. Occupations followed the Australian and New Zealand Standard Classification of Occupations (ANZSCO) [13] with the exception of the following responses which were collapsed into a single group: technician and trade, machinery operators and drivers, and labourers. Industries were coded using the Australian and New Zealand Standard Industrial Classification (ANZSIC) [14]. Other than the four most prevalent industries in our sample, the remaining industries were collapsed to form a single group of ‘other industries’. Detail of working status and location of work was collected during the third survey, reflecting current work conditions.

Current self-rated health was assessed with the first question from the Short Form 12-item health questionnaire [15], and psychological distress was determined using the 6-item Kessler Psychological Distress scale [16] where moderate-high distress was defined as scores of 11 or greater [17]. Pre-existing medical condition categories were collected at the baseline survey and coded as previously described [11]. Current financial stress was determined with the question ‘*What do you feel is the level of your financial stress today, on a scale of 1 to 10 where 1 is not at all stressed and 10 is as stressed as can be?*’, where responses were dichotomised into groups of responses greater that five, defined as experiencing financial stress, and responses less than or equal to five as no or limited financial stress. Financial resources were evaluated with the question ‘*If all of a sudden you had to get $2000 for something important, could the money be obtained within a week*?’ [18]. Responses of ‘*yes’* were categorised as having more financial resources and responses of ‘*no’* and ‘*don’t know*’ as having fewer financial resources. The degree of social interaction was evaluated using items from the Duke Social Support Index [19], which described social interactions during the week prior. Numerical responses of interaction encounters were dichotomised, where low levels of interactions were defined as: having spent no time in-person with any non-household members, attending no meetings, or as having telephone or online calls with fewer than one person each day on average. Reference groups are selections for clarity of interpretation, prioritised for (pseudo-)ordinal boundary values, and collapsed categories. The reference groups for gender and age group describe the average Australian worker.

### Analytical approach

For each participant, the binary yes/no responses to the question on workplaces reopening (Table 1) was converted to a 0 (no) or 1 (yes) score. The total number of concerns reported were summed as a score for the overall level of concern. Additionally, items were categorised into two sub-scores, reflecting two distinct groupings of concerns being infection risk and impacts on work and life at home.

Three scores were defined as study outcomes:

- *Overall concern*: A score for the total number concerns with a range of 0-14.
- *Infection risk*: A score describing the sum of a subset of items regarding the concern of coronavirus infection, and associated risk factors, with a range of 0-8.
- *Impact on work and home life*: A score describing a sum of a subset of items relating to impacts on working conditions (excluding changes associated with infection risk) and life at home with a range of 0-6.

Analyses focused on describing which concerns were more or less common for workers, and elucidating factors associated with particular concerns. The number and percentage of workers reporting concerns were calculated for each outcome (Table 1). In addition, the median, lower quartile (LQ) and upper quartile (UQ) were calculated for each outcome.

Negative binomial regression models were used to explore associations between independent variables and outcomes, with maximum likelihood estimates of the distributions’ parameters. Three regression models were calculated, one for each outcome. Models were exploratory, investigating the role of several independent variables on outcomes. Statistical tests used significance levels of 0.05 for all adjusted incident rate ratios (IRRs), alongside their corresponding 95% confidence interval (CI).

Additionally, to explore associations between independent variables and individual items in the fourteen-item list of concerns, we calculated binary regression models using each individual item (i.e., each concern) as dependent variables (Supplementary Table 1). Independent variables were consistent across all regression models. Statistical tests used significance levels of 0.05 for all adjusted odds ratios (ORs), alongside their corresponding 95% confidence interval (CI).

## Results

A total of 1383 participants completed the study survey. There were 214 unemployed participants that were not asked questions about workplaces reopening and thus were not included in the analyses, leaving a sample of 1169 eligible participants. A further 24 participants were excluded due to missing data for dependent variables, and 82 participants were excluded from regression models due to missing data for at least one dependent variable. In total, 1063 participants with complete data were included in regression analyses.

Among 1063 workers, 917 (86.3%) reported at least one concern about workplaces reopening (Table 1). A total of 876 (82.4%) participants reported concerns related to infection risk, while 568 (53.4%) reported concerns related to impact on work and home life. Overall, workers reported a median of 4 concerns (out of 14), composed of 3 regarding infection risk and 1 regarding impacts on work and home life.

The most common concerns were getting infected with COVID-19, infecting other people, being able to maintain physical distance from colleagues and being concerned about work colleagues coming to work when sick. The most common concerns related to work and home life was a reduction in the time able to be spent with family and friends, and concern about how changes in the workplace will impact ability to perform the job role. In contrast, access to hand sanitiser and losing the ability to work from home were concerns that were least commonly reported (Table 1).

**Table 2.** **Workplace reopening concerns for infection risk and changes to work and home life.**

|  | Cohort<br>N (%) | Adjusted incident rate ratio (IRR) [95% Confidence Interval] |  |  |
| --- | --- | --- | --- | --- |
|  |  | Overall concern | Infection risk | Impact on work and home life |
| Median number of concerns<sup>a</sup><br>(Lower – Upper Quartile) |  | 4 (2-6) | 3 (1-5) | 1 (0-2) |
| **Demographics and residence** |  |  |  |  |
| **Gender** |  |  |  |  |
| Female | 616 (57.9) | 1.01 [0.91, 1.13] | 0.94 [0.85, 1.05] | **1.24\*\*** [1.07, 1.43] |
| Male | 447 (42.1) | 1.00 (ref.) | 1.00 (ref.) | 1.00 (ref.) |
| **Age Group** |  |  |  |  |
| 18-24 years | 73 (6.9) | **1.29\*** [1.02, 1.63] | **1.28\*** [1.01, 1.61] | 1.29 [0.95, 1.75] |
| 25-34 years | 158 (14.9) | 1.09 [0.92, 1.28] | 1.09 [0.92, 1.29] | 1.08 [0.87, 1.35] |
| 35-44 years | 201 (18.9) | 1.00 (ref.) | 1.00 (ref.) | 1.00 (ref.) |
| 45-54 years | 284 (26.7) | 0.95 [0.83, 1.10] | 0.98 [0.85, 1.13] | 0.89 [0.74, 1.08] |
| 55-64 years | 280 (26.3) | 1.05 [0.90, 1.22] | 1.08 [0.93, 1.26] | 0.95 [0.77, 1.17] |
| 65+ years | 67 (6.3) | 0.86 [0.68, 1.09] | 0.89 [0.70, 1.13] | 0.76 [0.54, 1.09] |
| **Education** |  |  |  |  |
| High School (not completed) | 78 (7.3) | 0.97 [0.78, 1.21] | 1.03 [0.83, 1.29] | 0.81 [0.60, 1.11] |
| High School (completed) | 146 (13.7) | 0.96 [0.80, 1.15] | 1.02 [0.85, 1.22] | **0.77\*** [0.60, 0.98] |
| TAFE / Trade certificate | 296 (27.8) | 0.97 [0.84, 1.12] | 1.01 [0.87, 1.17] | 0.84<sup>f</sup> [0.70, 1.02] |
| University – undergraduate degree | 298 (28.0) | 0.99 [0.87, 1.13] | 1.06 [0.92, 1.21] | 0.85<sup>†</sup> [0.72, 1.02] |
| University – postgraduate degree | 245 (23.0) | 1.00 (ref.) | 1.00 (ref.) | 1.00 (ref.) |
| **Household situation** |  |  |  |  |
| Partner/spouse, no dependent children | 351 (33.0) | 1.14<sup>†</sup> [0.98, 1.33] | 1.09 [0.94, 1.27] | **1.36\*\*** [1.10, 1.69] |
| Partner/spouse, with dependent children | 326 (30.7) | 1.10 [0.94, 1.29] | 1.01 [0.87, 1.19] | **1.44\*\*** [1.16, 1.79] |
| Single parent with dependent children | 44 (4.1) | 1.06 [0.82, 1.38] | 0.97 [0.74, 1.27] | 1.41<sup>†</sup> [0.99, 2.01] |
| Other family members | 130 (12.2) | 1.06 [0.88, 1.29] | 1.03 [0.85, 1.26] | 1.22 [0.93, 1.59] |
| Other non-family members | 62 (5.8) | 1.10 [0.87, 1.38] | 1.05 [0.83, 1.33] | 1.26 [0.91, 1.73] |
| I live alone | 150 (14.1) | 1.00 (ref.) | 1.00 (ref.) | 1.00 (ref.) |
| **Residential Location** |  |  |  |  |
| Victoria (during an outbreak) | 386 (36.3) | 1.10<sup>†</sup> [0.99, 1.23] | 1.10<sup>†</sup> [0.98, 1.22] | 1.11 [0.96, 1.28] |
| Rest of Australia | 677 (63.7) | 1.00 (ref.) | 1.00 (ref.) | 1.00 (ref.) |
| **Work** |  |  |  |  |
| **Occupation** |  |  |  |  |
| Managers | 154 (14.5) | 0.86<sup>†</sup> [0.72, 1.02] | 0.87 [0.73, 1.04] | 0.87 [0.68, 1.11] |
| Professionals | 376 (35.4) | 1.02 [0.87, 1.20] | 1.02 [0.87, 1.20] | 1.03 [0.83, 1.28] |
| Community and Personal Service Workers | 126 (11.9) | 1.03 [0.86, 1.25] | 1.00 [0.83, 1.21] | 1.14 [0.89, 1.47] |
| Clerical and Administrative Workers | 134 (12.6) | 0.95 [0.80, 1.14] | 0.96 [0.80, 1.15] | 0.92 [0.72, 1.19] |
| Sales Workers | 72 (6.8) | 0.93 [0.73, 1.18] | 0.89 [0.70, 1.13] | 1.07 [0.78, 1.47] |
| Technician and Trade, Machinery Operators and Drivers, Labourers | 201 (18.9) | 1.00 (ref.) | 1.00 (ref.) | 1.00 (ref.) |
| **Industry** |  |  |  |  |
| Health Care and Social Assistance | 217 (20.4) | 1.14<sup>†</sup> [0.99, 1.30] | **1.16\*** [1.01, 1.33] | 1.09 [0.91, 1.31] |
| Education and Training | 138 (13.0) | 1.09 [0.93, 1.27] | 1.14 [0.97, 1.33] | 1.00 [0.81, 1.23] |
| Retail Trade | 83 (7.8) | **1.27\*** [1.03, 1.57] | **1.31\*** [1.06, 1.61] | 1.17 [0.88, 1.55] |
| Accommodation and Food Services | 62 (5.8) | 1.24<sup>†</sup> [1.00, 1.53] | **1.25\*** [1.01, 1.55] | 1.22 [0.92, 1.61] |
| Other Industries | 563 (53.0) | 1.00 (ref.) | 1.00 (ref.) | 1.00 (ref.) |
| **Employment type** |  |  |  |  |
| Casual | 233 (21.9) | 0.96 [0.85, 1.09] | 0.99 [0.86, 1.12] | 0.91 [0.77, 1.09] |
| Part-time | 219 (20.6) | 0.92 [0.81, 1.05] | 0.96 [0.84, 1.10] | **0.79\*** [0.66, 0.94] |
| Other (e.g. partner, contractor) | 15 (1.4) | 0.92 [0.62, 1.38] | 0.87 [0.58, 1.33] | 1.08 [0.66, 1.76] |
| Full-time | 596 (56.1) | 1.00 (ref.) | 1.00 (ref.) | 1.00 (ref.) |
| **Current work status** |  |  |  |  |
| Not Working | 115 (10.8) | **0.79\*\*** [0.66, 0.94] | **0.72\*\*** [0.60, 0.86] | 0.99 [0.80, 1.23] |
| Usual workplace | 618 (58.1) | **0.66\*\*** [0.58, 0.75] | **0.68\*\*** [0.60, 0.78] | **0.61\*\*** [0.52, 0.72] |
| Both working from home and usual workplace | 115 (10.8) | **0.76\*\*** [0.64, 0.91] | **0.75\*\*** [0.62, 0.89] | 0.81<sup>†</sup> [0.64, 1.02] |
| Working from home | 215 (20.2) | 1.00 (ref.) | 1.00 (ref.) | 1.00 (ref.) |
| **Health** |  |  |  |  |
| **Pre-existing medical conditions** |  |  |  |  |
| None | 653 (61.4) | 1.00 (ref.) | 1.00 (ref.) | 1.00 (ref.) |
| One | 232 (21.8) | 1.03 [0.91, 1.17] | 1.06 [0.93, 1.20] | 0.96 [0.81, 1.13] |
| Two or more | 178 (16.7) | 1.04 [0.88, 1.22] | 1.06 [0.90, 1.25] | 0.94 [0.76, 1.17] |
| **Psychological distress** |  |  |  |  |
| Low to None | 669 (62.9) | 1.00 (ref.) | 1.00 (ref.) | 1.00 (ref.) |
| High to Moderate | 394 (37.1) | **1.37\*\*** [1.23, 1.52] | **1.28\*\*** [1.14, 1.43] | **1.64\*\*** [1.41, 1.89] |
| **Self-rated health** |  |  |  |  |
| Poor to Good | 444 (41.8) | 1.06 [0.96, 1.17] | 1.04 [0.94, 1.15] | 1.13<sup>†</sup> [0.98, 1.29] |
| Very Good to Excellent | 619 (58.2) | 1.00 (ref.) | 1.00 (ref.) | 1.00 (ref.) |
| **Finance** |  |  |  |  |
| **Financial stress** |  |  |  |  |
| Yes | 282 (26.5) | **1.18\*\*** [1.05, 1.33] | 1.12<sup>†</sup> [1.00, 1.26] | **1.34\*\*** [1.15, 1.57] |
| No | 781 (73.5) | 1.00 (ref.) | 1.00 (ref.) | 1.00 (ref.) |
| **Financial resources** |  |  |  |  |
| No | 130 (12.2) | 1.04 [0.89, 1.21] | 1.00 [0.85, 1.17] | 1.19<sup>†</sup> [0.98, 1.46] |
| Yes | 933 (87.8) | 1.00 (ref.) | 1.00 (ref.) | 1.00 (ref.) |
| **Social interactions** |  |  |  |  |
| **Occasions spending time with others in-person** |  |  |  |  |
| None | 309 (29.1) | 1.04 [0.93, 1.15] | 1.04 [0.93, 1.16] | 1.07 [0.92, 1.23] |
| At least once | 754 (70.9) | 1.00 (ref.) | 1.00 (ref.) | 1.00 (ref.) |
| ***Meetings attended*** |  |  |  |  |
| None | 303 (28.5) | 0.94 [0.85, 1.05] | 0.93 [0.83, 1.03] | 1.01 [0.88, 1.17] |
| At least once | 760 (71.5) | 1.00 (ref.) | 1.00 (ref.) | 1.00 (ref.) |
| ***Made or received telephone / online calls*** |  |  |  |  |
| Fewer than seven occasions | 423 (39.8) | 0.99 [0.90, 1.09] | 0.99 [0.90, 1.10] | 0.99 [0.87, 1.13] |
| Seven or more occasions | 640 (60.2) | 1.00 (ref.) | 1.00 (ref.) | 1.00 (ref.) |
| **Survey mode** |  |  |  |  |
| Online | 328 (30.9) | 0.88<sup>†</sup> [0.76, 1.01] | 0.93 [0.81, 1.07] | **0.74\*\*** [0.61, 0.89] |
| Telephone | 735 (69.1) | 1.00 (ref.) | 1.00 (ref.) | 1.00 (ref.) |

<sup>†</sup>0.05≤p<0.1, \*0.01≤p<0.05, \*\*p<0.01. <sup>a</sup>maximum scores for overall concerns, infection risk, and impacts on work and home life are 14, 8, and 6 respectively.

### Overall infection risk

Risks of infection were reported as a concern for 82.4% of workers. Participants reported a significantly greater number of concerns regarding infection risk if they were aged 18-24 years (IRR=1.28, 95% CI=[1.01, 1.61]), working in the health care and social assistance (IRR=1.16, 95% CI=[1.01, 1.33]), retail (IRR=1.31, 95% CI=[1.06, 1.61]), and the accommodation and food services industry (IRR=1.25, 95% CI=[1.01, 1.55]) (Table 2). Compared to participants working exclusively from home, significantly fewer concerns regarding infection risks were reported by participants working at their usual workplace (IRR=0.68, 95% CI=[0.60, 0.78]), working both at their usual workplace and at home (IRR=0.75, 95% CI=[0.62, 0.89]), or those employed but not working (IRR=0.72, 95% CI=[0.60, 0.86]). Workers reporting psychological distress also reported a significantly larger number of concerns (IRR=1.28, 95% CI=[1.13, 1.43]), as did workers reporting financial stress (IRR=1.12, 95% CI=[1.00, 1.26]).

### Specific infection risks

Parents with dependent children were more worried about getting infected, as were health care, retail, and Victorian workers (Supplementary Table 1). The worry of infecting others was less commonly reported by managers, and part-time workers compared to reference groups. Distancing from members of the public was a common concern for workers across industries such as health care, retail, accommodation and food sectors. Distancing from colleagues was a worry of health care workers, people working in education and workers in Victoria (Supplementary Table 1). Access to soap and sanitisers was a less common worry for female workers, managers, and administration staff (Supplementary Table 1).

### Overall impacts on work and home life

Concerns relating to changes in work and home life were reported by 53.4% of participants. Female workers reported significantly more concerns about changes to work and home life (IRR=1.24, 95% CI=[1.07, 1.43]), as did workers living with a spouse and either with (IRR=1.36, 95% CI=[1.10, 1.69]) or without (IRR=1.44, 95% CI=[1.16, 1.79]) dependent children (Table 2). Participants whose highest level of education was completing high school had significantly fewer concerns (IRR=0.77, 95% CI=[0.60, 0.98]) than those with a post-graduate university degree. Fewer concerns about the impact on work and home life were reported by participants working at their usual workplace (IRR=0.61, 95% CI=[0.52, 0.72]), and among part-time workers (IRR=0.79, 95% CI=[0.66,0.94]). Participants experiencing psychological distress reported more concerns about the impact on work and home life (IRR=1.64, 95% CI=[1.41, 1.89]), as did those in financial stress (IRR=1.34, 95% CI=[1.15, 1.57]).

### Specific impacts on home and work life

Workers most concerned about how changes to the workplace would affect their ability to work were people in accommodation or the food services industry, as were people that were employed but were temporarily not working (Supplementary Table 1). Uncertainty of job role was a common concern for Victorian workers, community and personal service and people currently employed but not working (Supplementary Table 1). Losing the ability to work from home was highlighted by workers experiencing financial stress. People that were working casually or part-time were less concerned about losing the ability to work from home. Changes in roles and responsibilities at home were more common concerns reported by single parents, couples living together either with or without children, people living with other family members, and individuals in financial stress (Supplementary Table 1).

## Discussion

The majority of Australian workers enrolled in this study reported concerns about returning to the workplace during the COVID-19 pandemic. Data was collected in the third quarter of 2020 while Australia’s second largest state was experiencing a substantial second wave of COVID-19 cases with an associated extended community lockdown and business closures, while the rest of the nation was gradually re-opening. More than four out of five workers expressed concerns about infection risk in the workplace, while slightly more than half reported at least one concern about the impacts of workplaces reopening on their work and home life. Our findings also demonstrate that the prevalence of reported concerns varies according to some occupational, demographic, health and social characteristics of workers.

Workers in some industries have an inherent higher risk of occupational infection [4], for example workers in healthcare, aged care, food services and the retail trade where a large proportion of the workforce regularly interact in close proximity to the public. Our analysis suggests that workers’ concerns about infection risks are consistent with the reported high rates of transmission in these industries. The nature of these concerns also appeared to be related to the types of occupational exposure experienced in these industries. For example, healthcare workers report being worried about being infected, infecting others and maintaining physical distancing, but not about hygiene practices that are commonly addressed in these settings such as handwashing.

Independent of occupational factors, concerns about reopening are also related to demographic, health and social factors such as worker’s age, their psychological and financial state, and their household composition. Concerns regarding pressures to keep working whilst sick were more common among younger workers in our analysis. This may, in part, reflect some of the insecurities and lack of paid sick leave for workers in casual employment or the gig economy, in which a high proportion of workers are people aged under twenty-five [20] and consistent with fewer supports for casual workers internationally [21]. Whist the implementation of pandemic leave payments provides financial support to discourage people without leave entitlements working whilst sick [22-24], these actions may not be enough to encourage all sick workers to isolate from others, and thus alleviate the perceived risk of people working whilst sick.

People who were working from home were more likely to report more concerns than those who had either maintained some time at work, or whose work had ceased despite remaining employed. This may indicate a lack of exposure or lesser understanding of policies and procedures introduced to minimise workplace infections, resulting in heightened anxiety about returning to the workplace. Alternatively, or additionally, those maintaining some engagement in the workplace may have realised fewer of the benefits of working from home, and thus be less concerned about any impact of returning on home and work life.

Household circumstances were statistically related to reported concerns. Workers living with family members reported more concerns of both infection risk and impacts on home and work life. These findings suggest a desire to maintain the benefits of working from home arrangements, and potentially concerns about workplace infections being transmitted into the home environment. Our data suggests that people with additional home-based responsibilities, such as parents with dependent children, will be more likely to report concerns of returning to the workplaces on their home lives. We also observe that workers reporting moderate to high levels of psychological distress are more likely to express concerns about returning to the workplace. This effect is larger for concerns related to impacts on home and work life, but is also present for concerns related to risk of infection in the workplace. We and others have previously reported on the high prevalence of psychological distress [11] and mental health concerns [25] among workers during the COVID-19 pandemic. These workers may warrant particular attention by employers seeking to re-engage their workforces in the physical work environment. Programs that can support identification of workers with distress, and then provision of effective mental health support, may alleviate workers concerns. Strategies that alleviate concerns about returning to the workplace may also support reductions in psychological distress.

To our knowledge, this is one of the first studies reporting the concerns of workers regarding returning to the workplaces during the COVID-19 pandemic. Study strengths include a large, national sample, use of standardised measures to assess co-variates such as health, occupation and industry and the ability to assess the relationship between multiple confounders in multivariate regression models. The sample may not be representative of the national working age population, although the regression model adjusts for multiple demographic factors. The cross-sectional nature of the study limits causal interpretation. Data collection occurred during the second wave of COVID-19 cases in Australia [8] allowing the comparison of workers within the state of this outbreak with those outside, providing insight into the role played by the degree of community transmission on concerns of workers such as an increased level of uncertainty in job roles upon returning to the workplace and distancing from co-workers.

Worker concerns are likely to evolve over time as infection risk and risk perceptions change. For example, the progressive vaccination for SARS-CoV-2 currently underway in many nations may influence the concerns of workers. Worker concerns may change as vaccination rates rise and as efficacy is better understood in the workplace, community or national settings. Emergence of viral variants that are resistant to vaccination may also affect worker concerns. Our findings suggest a need for follow-on studies of the study barriers and facilitators to returning to workplaces, to develop a stronger evidence base to support workers and employers in future.

Although we are not aware of any definitive data, it appears that most organisations have modified the physical workplace environment to reduce the risk of viral transmission, while many governments have mandated such changes. These measures will address some, but not all, of the concerns workers express regarding returning to the workplace. Our findings indicate clearly that employers must also address concerns related to impacts on home life, family and social impacts and job roles. Given the high rate of concerns related to the physical work environment, it seems clear that effective communication of infection control measures to returning workers will support transitions back into the workplace. Responses should be tailored to industrial and workforce characteristics – for example a greater focus on interactions with other staff and members of the public in health and aged care settings, and a greater focus on home and work life impacts on people with partners and children at home. Organisations that are able to maintain working from home arrangements may be able to reduce the concerns regarding impacts on home and work life by preserving (wholly or partly) remote working conditions, or providing greater flexibility in working arrangements such as modified hours. The importance of a credible reduction of infection risk and adequate workplace hygiene is fundamental to mitigate the concerns of workers, otherwise predisposed concerned workers and members of the public may continue to self-impose public health measures after they are eased [26].

## Conclusion

The COVID-19 pandemic has resulted in dynamic and widespread changes in employment globally. In this study of Australian workers, most workers reported concerns about workplaces reopening. The most prevalent concerns related to workplace infection risk, but a slight majority of workers also reported concerns related to changes in work and impacts on life at home. The concerns of workers are not shared equally – occupational, personal and family circumstances are related to the nature and prevalence of concerns. Actions alleviating concerns of workers are recommended. This should include policies and procedures that minimise infection risk in the workplace, with considerations given to interactions with other people, workplace hygiene, and the use of public transportation. Organisational changes should be reinforced and clearly communicated with workers. Changes to policies on remote working so that workers can continue to work from home, at least in part, or to have more flexible working arrangements, may also reduce concerns related to impacts to life at home. The concerns of workers are likely to change as the pandemic unfolds, and thus further research monitoring these issues over time will inform future evidence-based workplace policy recommendations.

## Data Availability

The data are held at Monash University, Insurance Work and Health Group, School of Public Health and Preventive Medicine. Procedures to access data from this study are available through contacting the lead author. Proposals for collaborative analyses will be considered by the study's investigator team.

## Notes

### Competing Interest Statement

Funding was provided by Monash University and the icare Foundation. The views expressed are those of the authors and may not reflect the views of study funders.

### Clinical Trial

ACTRN12620000857909

### Funding Statement

Funding was provided by Monash University and the icare Foundation. The views expressed are those of the authors and may not reflect the views of study funders. Professor Alex Collie is supported by an ARC Future Fellowship.

### Author Declarations

Approval to conduct the study was provided by Monash University Human Research Ethics Committee.

