## Supplemental Table 1 for "Returning to the workplace during the COVID-19 pandemic: The concerns of Australian workers"

### Supplementary Information

#### Table S1. Adjusted binary regression models describing fourteen items of concerns about workplaces reopening.

| Adjusted Odds Ratio (OR) [95% Confidence Interval] | | | | | | | |
|---|---|---|---|---|---|---|---|
| **Concern** | Getting infected with COVID-19 | Work colleagues coming to work while sick | Infecting someone else with COVID-19 | Maintaining physical distance from my work colleagues | Maintaining physical distance from members of the public | Appropriate cleaning of my workplace | Travelling to and from work on public transport |
| **Workers with concern N (%)** | **525 (49.4)** | **521 (49.0)** | **520 (48.9)** | **488 (45.9)** | **365 (34.3)** | **311 (29.3)** | **253 (23.8)** |
| **Demographics and residence** | | | | | | | |
| ***Gender*** | | | | | | | |
| Female | 1.26 [0.94, 1.70] | 0.88 [0.66, 1.19] | 1.30† [0.96, 1.76] | 0.87 [0.63, 1.19] | 0.81 [0.60, 1.10] | 0.77 [0.55, 1.07] | **0.68*** [0.48, 0.97] |
| Male | 1.00 (ref.) | 1.00 (ref.) | 1.00 (ref.) | 1.00 (ref.) | 1.00 (ref.) | 1.00 (ref.) | 1.00 (ref.) |
| ***Age Group*** | | | | | | | |
| 18-24 year | 1.38 [0.71, 2.66] | 1.90† [0.98, 3.69] | 1.37 [0.70, 2.68] | 1.76 [0.87, 3.54] | 1.27 [0.65, 2.48] | 1.94† [0.95, 3.97] | 1.31 [0.60, 2.86] |
| 25-34 year | 1.23 [0.77, 1.96] | 1.01 [0.63, 1.61] | 1.20 [0.75, 1.93] | 1.28 [0.78, 2.12] | 1.38 [0.86, 2.22] | 1.39 [0.83, 2.35] | 0.98 [0.57, 1.68] |
| 35-44 years | 1.00 (ref.) | 1.00 (ref.) | 1.00 (ref.) | 1.00 (ref.) | 1.00 (ref.) | 1.00 (ref.) | 1.00 (ref.) |
| 45-54 years | 1.01 [0.69, 1.49] | 0.90 [0.61, 1.32] | 0.90 [0.61, 1.33] | 1.16 [0.76, 1.76] | 0.94 [0.63, 1.38] | 1.30 [0.84, 2.02] | 0.65† [0.41, 1.04] |
| 55-64 years | 1.61† [1.06, 2.44] | 0.83 [0.55, 1.26] | 1.14 [0.75, 1.75] | 1.32 [0.84, 2.07] | 1.08 [0.71, 1.65] | 1.49† [0.93, 2.39] | 0.81 [0.49, 1.34] |
| 65+ year | 1.43 [0.76, 2.67] | **0.52*** [0.27, 1.00] | 0.63 [0.33, 1.21] | 1.54 [0.79, 3.02] | 0.92 [0.49, 1.75] | 0.94 [0.44, 1.98] | **0.38*** [0.16, 0.91] |
| ***Education*** | | | | | | | |
| High School (not completed) | 1.56 [0.86, 2.85] | 1.35 [0.74, 2.45] | 0.90 [0.49, 1.66] | 1.19 [0.61, 2.30] | 1.38 [0.75, 2.52] | 0.86 [0.43, 1.72] | **0.42*** [0.18, 0.98] |
| High School (completed) | 0.97 [0.60, 1.59] | 1.25 [0.77, 2.04] | 1.03 [0.63, 1.70] | 1.15 [0.68, 1.96] | 1.04 [0.63, 1.73] | 1.06 [0.62, 1.80] | 0.58† [0.32, 1.04] |
| TAFE / Trade certificate | 1.12 [0.75, 1.67] | 1.16 [0.78, 1.72] | 0.94 [0.63, 1.40] | 1.23 [0.81, 1.89] | 1.17 [0.78, 1.75] | 0.85 [0.55, 1.32] | **0.54*** [0.33, 0.86] |
| University – undergraduate degree | 1.07 [0.74, 1.53] | 1.17 [0.81, 1.69] | 0.94 [0.65, 1.36] | **1.48*** [1.00, 2.19] | 1.11 [0.77, 1.61] | 1.04 [0.70, 1.56] | 0.89 [0.59, 1.35] |
| University – postgraduate degree | 1.00 (ref.) | 1.00 (ref.) | 1.00 (ref.) | 1.00 (ref.) | 1.00 (ref.) | 1.00 (ref.) | 1.00 (ref.) |
| ***Household situation*** | | | | | | | |
| Partner/spouse, no dependent children | 1.45† [0.96, 2.18] | 1.15 [0.76, 1.73] | 1.26 [0.82, 1.91] | 1.18 [0.76, 1.82] | 1.44† [0.95, 2.18] | 0.99 [0.64, 1.55] | 1.30 [0.79, 2.15] |
| Partner/spouse, with dependent children | **1.62*** [1.06, 2.49] | 1.00 [0.65, 1.52] | 1.09 [0.71, 1.69] | 1.15 [0.73, 1.82] | 1.15 [0.75, 1.78] | 0.92 [0.58, 1.47] | 0.76 [0.45, 1.28] |

| | | | | | | | |
|---|---|---|---|---|---|---|---|
| Single parent with dependent children | 1.38 [0.66, 2.89] | 0.81 [0.39, 1.66] | 1.09 [0.52, 2.30] | 1.78 [0.83, 3.84] | 1.47 [0.71, 3.08] | 0.77 [0.34, 1.78] | 0.60 [0.23, 1.55] |
| Other family members | 1.44 [0.84, 2.47] | **0.84** [0.49, 1.43] | 1.70† [0.98, 2.95] | 1.13 [0.64, 2.01] | 1.50 [0.88, 2.59] | 0.90 [0.50, 1.62] | 0.93 [0.48, 1.83] |
| Other non-family members | 1.51 [0.79, 2.91] | 1.02 [0.53, 1.94] | 1.10 [0.57, 2.14] | 1.89† [0.97, 3.71] | 1.02 [0.53, 1.97] | 0.83 [0.41, 1.68] | 1.01 [0.47, 2.20] |
| I live alone | 1.00 (ref.) | 1.00 (ref.) | 1.00 (ref.) | 1.00 (ref.) | 1.00 (ref.) | 1.00 (ref.) | 1.00 (ref.) |
| ***Residential Location*** | | | | | | | |
| Victoria (during an outbreak) | **1.57*** [1.16, 2.12] | 0.95 [0.71, 1.28] | 1.24 [0.92, 1.68] | **1.53**** [1.12, 2.09] | 1.10 [0.81, 1.48] | 1.17 [0.85, 1.63] | 0.80 [0.55, 1.16] |
| Rest of Australia | 1.00 (ref.) | 1.00 (ref.) | 1.00 (ref.) | 1.00 (ref.) | 1.00 (ref.) | 1.00 (ref.) | 1.00 (ref.) |
| **Work** | | | | | | | |
| ***Occupation*** | | | | | | | |
| Managers | 0.72 [0.45, 1.16] | 0.98 [0.62, 1.56] | **0.53*** [0.33, 0.86] | 1.05 [0.64, 1.75] | 0.81 [0.50, 1.31] | 0.73 [0.43, 1.25] | 1.52 [0.83, 2.76] |
| Professionals | 0.95 [0.61, 1.47] | 0.96 [0.62, 1.48] | 0.90 [0.58, 1.40] | 0.97 [0.61, 1.54] | 1.23 [0.79, 1.91] | 0.92 [0.57, 1.49] | **2.43**** [1.40, 4.22] |
| Community and Personal Service Workers | 1.07 [0.64, 1.78] | 1.15 [0.69, 1.90] | 0.70 [0.41, 1.18] | 0.74 [0.42, 1.28] | 1.38 [0.83, 2.32] | 0.96 [0.55, 1.67] | 1.11 [0.53, 2.29] |
| Clerical and Administrative Workers | 0.93 [0.56, 1.52] | 0.93 [0.57, 1.53] | 0.99 [0.60, 1.64] | 0.93 [0.54, 1.58] | 0.85 [0.51, 1.42] | 0.94 [0.54, 1.63] | **1.73**** [0.91, 3.27] |
| Sales Workers | 0.56† [0.28, 1.08] | 0.92 [0.48, 1.78] | 0.75 [0.38, 1.45] | 0.83 [0.41, 1.69] | 0.99 [0.50, 1.94] | 0.77 [0.37, 1.59] | 1.40 [0.63, 3.10] |
| Technician and Trade, Machinery Operators and Drivers, Labourers | 1.00 (ref.) | 1.00 (ref.) | 1.00 (ref.) | 1.00 (ref.) | 1.00 (ref.) | 1.00 (ref.) | 1.00 (ref.) |
| ***Industry*** | | | | | | | |
| Health Care and Social Assistance | **1.69**** [1.16, 2.46] | 0.95 [0.65, 1.38] | **1.62*** [1.10, 2.37] | **1.56*** [1.05, 2.33] | **1.84**** [1.26, 2.68] | 1.25 [0.82, 1.89] | 0.72 [0.45, 1.16] |
| Education and Training | 1.40 [0.91, 2.16] | 1.02 [0.66, 1.57] | 1.54† [0.99, 2.40] | **1.82*** [1.15, 2.86] | 1.23 [0.79, 1.90] | **1.81*** [1.14, 2.87] | **0.45**** [0.25, 0.80] |
| Retail Trade | **2.00*** [1.10, 3.62] | 0.87 [0.48, 1.55] | **1.90*** [1.05, 3.46] | 1.33 [0.72, 2.48] | **2.63**** [1.44, 4.79] | 1.65 [0.88, 3.10] | **1.94*** [0.99, 3.82] |
| Accommodation and Food Services | 1.77† [0.97, 3.24] | 0.87 [0.48, 1.59] | 1.72† [0.93, 3.18] | 1.50 [0.80, 2.82] | **2.93**** [1.57, 5.48] | 1.00 [0.52, 1.94] | 1.65 [0.80, 3.38] |
| Other Industries | 1.00 (ref.) | 1.00 (ref.) | 1.00 (ref.) | 1.00 (ref.) | 1.00 (ref.) | 1.00 (ref.) | 1.00 (ref.) |
| ***Employment type*** | | | | | | | |
| Casual | 0.88 [0.62, 1.26] | 1.09 [0.76, 1.55] | 0.77 [0.53, 1.11] | 1.09 [0.75, 1.60] | 1.01 [0.70, 1.45] | 1.03 [0.70, 1.53] | 1.01 [0.65, 1.58] |
| Part-time | 0.82 [0.57, 1.18] | 0.77 [0.54, 1.10] | **0.69*** [0.48, 0.99] | 1.21 [0.83, 1.77] | 1.14 [0.79, 1.64] | 1.13 [0.76, 1.68] | 0.80 [0.51, 1.27] |
| Other (e.g. partner, contractor) | 0.80 [0.26, 2.44] | 2.58 [0.82, 8.11] | 0.34† [0.11, 1.08] | 1.31 [0.42, 4.14] | 0.31† [0.08, 1.22] | 0.72 [0.19, 2.81] | 0.75 [0.19, 2.97] |
| Full-time | 1.00 (ref.) | 1.00 (ref.) | 1.00 (ref.) | 1.00 (ref.) | 1.00 (ref.) | 1.00 (ref.) | 1.00 (ref.) |
| ***Current work status*** | | | | | | | |
| Not Working | 0.63† [0.38, 1.05] | **0.48**** [0.29, 0.80] | 0.65 [0.39, 1.09] | **0.50*** [0.30, 0.85] | 0.71 [0.43, 1.18] | **0.47**** [0.28, 0.81] | **0.30**** [0.17, 0.54] |

| | | | | | | | |
|---|---|---|---|---|---|---|---|
| Usual workplace | **0.69*** [0.48, 1.00] | **0.36**** [0.25, 0.53] | 0.77 [0.53, 1.11] | **0.41**** [0.28, 0.60] | **0.62*** [0.43, 0.90] | **0.39**** [0.26, 0.57] | **0.24**** [0.16, 0.36] |
| Both working from home and usual workplace | 0.95 [0.58, 1.54] | **0.42**** [0.26, 0.68] | 0.82 [0.50, 1.35] | 0.61† [0.37, 1.02] | **0.47**** [0.28, 0.79] | **0.37**** [0.21, 0.65] | **0.41**** [0.24, 0.71] |
| Working from home | 1.00 (ref.) | 1.00 (ref.) | 1.00 (ref.) | 1.00 (ref.) | 1.00 (ref.) | 1.00 (ref.) | 1.00 (ref.) |
| **Health** | | | | | | | |
| ***Pre-existing medical conditions*** | | | | | | | |
| None | 1.00 (ref.) | 1.00 (ref.) | 1.00 (ref.) | 1.00 (ref.) | 1.00 (ref.) | 1.00 (ref.) | 1.00 (ref.) |
| One | 1.06 [0.75, 1.50] | 1.14 [0.81, 1.61] | 1.09 [0.76, 1.55] | 1.34 [0.93, 1.93] | 1.10 [0.77, 1.56] | 1.08 [0.74, 1.57] | 0.77 [0.50, 1.17] |
| Two or more | 1.25 [0.80, 2.00] | 1.40 [0.90, 2.18] | 1.08 [0.68, 1.71] | 1.55† [0.97, 2.47] | 1.25 [0.80, 1.96] | 0.82 [0.51, 1.33] | 0.61† [0.34, 1.09] |
| ***Psychological distress*** | | | | | | | |
| Low to None | 1.00 (ref.) | 1.00 (ref.) | 1.00 (ref.) | 1.00 (ref.) | 1.00 (ref.) | 1.00 (ref.) | 1.00 (ref.) |
| High to Moderate | **1.69**** [1.25, 2.30] | **1.42*** [1.05, 1.92] | **1.84**** [1.34, 2.52] | **1.72**** [1.25, 2.36] | **1.58**** [1.17, 2.14] | **1.65**** [1.19, 2.29] | **1.49*** [1.03, 2.16] |
| ***Self-rated health*** | | | | | | | |
| Poor to Good | 1.17 [0.89, 1.54] | 0.98 [0.75, 1.29] | 1.04 [0.78, 1.37] | 0.89 [0.66, 1.19] | 1.25 [0.95, 1.65] | 1.05 [0.78, 1.42] | 1.04 [0.74, 1.46] |
| Very Good to Excellent | 1.00 (ref.) | 1.00 (ref.) | 1.00 (ref.) | 1.00 (ref.) | 1.00 (ref.) | 1.00 (ref.) | 1.00 (ref.) |
| **Finance** | | | | | | | |
| ***Financial stress*** | | | | | | | |
| Yes | 1.34† [0.95, 1.87] | 1.28 [0.91, 1.78] | 1.02 [0.72, 1.44] | **1.46*** [1.03, 2.07] | 1.23 [0.87, 1.72] | 1.39† [0.97, 1.99] | 1.12 [0.74, 1.68] |
| No | 1.00 (ref.) | 1.00 (ref.) | 1.00 (ref.) | 1.00 (ref.) | 1.00 (ref.) | 1.00 (ref.) | 1.00 (ref.) |
| ***Financial resources*** | | | | | | | |
| No | 0.70 [0.45, 1.10] | 1.05 [0.67, 1.63] | 0.79 [0.50, 1.25] | 0.97 [0.61, 1.56] | 0.98 [0.62, 1.54] | 1.16 [0.72, 1.85] | 1.50 [0.87, 2.61] |
| Yes | 1.00 (ref.) | 1.00 (ref.) | 1.00 (ref.) | 1.00 (ref.) | 1.00 (ref.) | 1.00 (ref.) | 1.00 (ref.) |
| **Social interactions** | | | | | | | |
| ***Occasions spending time with others in-person*** | | | | | | | |
| None | 0.98 [0.72, 1.34] | 1.27 [0.94, 1.73] | 0.98 [0.72, 1.34] | 1.13 [0.82, 1.56] | 0.90 [0.66, 1.23] | 1.11 [0.79, 1.54] | 1.13 [0.78, 1.64] |
| At least once | 1.00 (ref.) | 1.00 (ref.) | 1.00 (ref.) | 1.00 (ref.) | 1.00 (ref.) | 1.00 (ref.) | 1.00 (ref.) |
| ***Meetings attended*** | | | | | | | |
| None | 0.80 [0.60, 1.08] | 1.02 [0.77, 1.37] | 0.80 [0.59, 1.07] | 0.85 [0.62, 1.17] | 0.91 [0.68, 1.22] | 0.94 [0.68, 1.30] | 0.86 [0.60, 1.24] |
| At least one | 1.00 (ref.) | 1.00 (ref.) | 1.00 (ref.) | 1.00 (ref.) | 1.00 (ref.) | 1.00 (ref.) | 1.00 (ref.) |
| ***Made or received telephone / online calls*** | | | | | | | |

| | | | | | | | |
|---|---|---|---|---|---|---|---|
| Fewer than seven occasions | 1.10 [0.84, 1.43] | 0.98 [0.76, 1.28] | 0.99 [0.75, 1.29] | 0.79 [0.60, 1.05] | 1.04 [0.79, 1.35] | 1.05 [0.79, 1.41] | 0.97 [0.70, 1.35] |
| Seven or more occasions | 1.00 (ref.) | 1.00 (ref.) | 1.00 (ref.) | 1.00 (ref.) | 1.00 (ref.) | 1.00 (ref.) | 1.00 (ref.) |
| **Survey mode** | | | | | | | |
| Online | 0.92 [0.63, 1.35] | 0.96 [0.65, 1.41] | **0.29**** [0.19, 0.43] | 0.95 [0.64, 1.42] | 1.09 [0.74, 1.60] | 1.47† [0.98, 2.22] | 1.04 [0.65, 1.67] |
| Telephone | 1.00 (ref.) | 1.00 (ref.) | 1.00 (ref.) | 1.00 (ref.) | 1.00 (ref.) | 1.00 (ref.) | 1.00 (ref.) |

†0.05≤p<0.1, *0.01≤p<0.05, **p<0.01.

**Table S1 (continued). Adjusted binary regression models describing fourteen items of concerns about workplaces reopening.**

| | Adjusted Odds Ratio (OR) [95% Confidence Interval] | | | | | | |
|---|---|---|---|---|---|---|---|
| **Concern** | Access to soap or hand sanitiser | Reduction in the time I can spend with family / friends. | How changes to the workplace will affect my ability to work | Uncertainty around what my job will involve when I go back | Impact on my roles and responsibilities at home | Pressure to continue working even when feeling sick | Losing the ability to work from home |
| **Workers with concern N (%)** | **135 (12.7)** | **251 (23.6)** | **244 (23.0)** | **185 (17.4)** | **168 (15.8)** | **156 (14.7)** | **131 (12.3)** |
| **Demographics and residence** | | | | | | | |
| ***Gender*** | | | | | | | |
| Female | **0.57*** [0.37, 0.89] | **1.47*** [1.03, 2.10] | 1.41† [0.98, 2.02] | 1.16 [0.77, 1.74] | 1.47† [0.97, 2.22] | 1.09 [0.71, 1.67] | 1.58† [0.97, 2.58] |
| Male | 1.00 (ref.) | 1.00 (ref.) | 1.00 (ref.) | 1.00 (ref.) | 1.00 (ref.) | 1.00 (ref.) | 1.00 (ref.) |
| ***Age Group*** | | | | | | | |
| 18-24 year | 2.03 [0.84, 4.93] | 1.11 [0.53, 2.34] | 1.38 [0.62, 3.05] | 1.31 [0.53, 3.26] | 1.41 [0.56, 3.58] | 2.03† [0.92, 4.45] | 1.76 [0.59, 5.21] |
| 25-34 year | 0.88 [0.44, 1.77] | 0.86 [0.51, 1.46] | 1.38 [0.78, 2.44] | 1.44 [0.77, 2.69] | 1.36 [0.74, 2.51] | 0.85 [0.46, 1.56] | 0.91 [0.44, 1.88] |
| 35-44 years | 1.00 (ref.) | 1.00 (ref.) | 1.00 (ref.) | 1.00 (ref.) | 1.00 (ref.) | 1.00 (ref.) | 1.00 (ref.) |
| 45-54 years | 0.98 [0.55, 1.76] | 0.70 [0.45, 1.09] | 1.44 [0.89, 2.34] | 0.94 [0.54, 1.62] | 0.89 [0.54, 1.47] | 0.60† [0.35, 1.02] | 0.78 [0.43, 1.41] |
| 55-64 years | 1.05 [0.56, 1.96] | 0.89 [0.55, 1.44] | 1.30 [0.76, 2.20] | 1.45 [0.80, 2.61] | 0.82 [0.46, 1.45] | 0.59† [0.33, 1.07] | 0.81 [0.42, 1.57] |
| 65+ year | 0.60 [0.20, 1.81] | 0.51 [0.22, 1.18] | 1.56 [0.71, 3.45] | 1.12 [0.44, 2.86] | 0.71 [0.26, 1.93] | 0.33† [0.10, 1.06] | 0.41 [0.11, 1.57] |
| ***Education*** | | | | | | | |
| High School (not completed) | 0.77 [0.30, 1.97] | 1.12 [0.55, 2.27] | 0.59 [0.29, 1.22] | 0.73 [0.32, 1.68] | 0.59 [0.25, 1.42] | 0.94 [0.37, 2.40] | 0.35 [0.07, 1.69] |
| High School (completed) | 1.12 [0.54, 2.31] | 0.77 [0.41, 1.43] | 0.61 [0.33, 1.10] | 0.55† [0.28, 1.07] | **0.39*** [0.19, 0.82] | 1.37 [0.69, 2.71] | 0.93 [0.43, 2.03] |
| TAFE / Trade certificate | 0.83 [0.45, 1.53] | 1.10 [0.69, 1.77] | **0.54**** [0.33, 0.86] | 0.60† [0.35, 1.03] | 0.60† [0.35, 1.02] | 1.42 [0.80, 2.52] | 0.72 [0.39, 1.31] |
| University – undergraduate degree | 1.40 [0.82, 2.41] | 1.37 [0.89, 2.10] | **0.50**** [0.32, 0.76] | 0.66† [0.41, 1.08] | 0.76 [0.47, 1.23] | 1.08 [0.62, 1.87] | 0.84 [0.49, 1.44] |
| University – postgraduate degree | 1.00 (ref.) | 1.00 (ref.) | 1.00 (ref.) | 1.00 (ref.) | 1.00 (ref.) | 1.00 (ref.) | 1.00 (ref.) |
| ***Household situation*** | | | | | | | |
| Partner/spouse, no dependent children | 0.85 [0.49, 1.49] | 1.04 [0.64, 1.69] | 1.42 [0.84, 2.39] | 1.62 [0.91, 2.90] | **3.60**** [1.71, 7.57] | 1.26 [0.67, 2.39] | 1.73 [0.87, 3.44] |
| Partner/spouse, with dependent children | 0.58† [0.32, 1.05] | 1.11 [0.67, 1.84] | 1.46 [0.85, 2.51] | 1.42 [0.77, 2.60] | **5.48**** [2.60, 11.54] | 1.56 [0.82, 2.97] | 1.46 [0.73, 2.92] |
| Single parent with dependent children | 0.53 [0.16, 1.69] | 1.59 [0.69, 3.70] | 1.68 [0.70, 4.04] | 1.01 [0.34, 3.03] | **5.62**** [2.00, 15.76] | 0.58 [0.16, 2.19] | 1.72 [0.58, 5.13] |
| Other family members | 0.46† [0.20, 1.03] | 1.00 [0.53, 1.87] | 1.58 [0.82, 3.04] | 0.97 [0.45, 2.07] | **2.59*** [1.06, 6.32] | 1.34 [0.62, 2.89] | 0.99 [0.37, 2.62] |

| | | | | | | | |
|---|---|---|---|---|---|---|---|
| Other non-family members | 0.67 [0.27, 1.64] | 0.77 [0.35, 1.71] | **2.69*** [1.27, 5.69] | 2.20† [0.97, 5.03] | 1.01 [0.30, 3.40] | 1.22 [0.50, 2.97] | 0.90 [0.28, 2.92] |
| I live alone | 1.00 (ref.) | 1.00 (ref.) | 1.00 (ref.) | 1.00 (ref.) | 1.00 (ref.) | 1.00 (ref.) | 1.00 (ref.) |
| ***Residential Location*** | | | | | | | |
| Victoria (during an outbreak) | 0.89 [0.57, 1.39] | 1.26 [0.89, 1.78] | 1.29 [0.90, 1.85] | **1.64*** [1.10, 2.45] | 1.36 [0.91, 2.04] | 0.67† [0.43, 1.03] | 0.71 [0.44, 1.17] |
| Rest of Australia | 1.00 (ref.) | 1.00 (ref.) | 1.00 (ref.) | 1.00 (ref.) | 1.00 (ref.) | 1.00 (ref.) | 1.00 (ref.) |
| **Work** | | | | | | | |
| ***Occupation*** | | | | | | | |
| Managers | **0.44*** [0.21, 0.91] | 0.61† [0.35, 1.09] | 1.31 [0.74, 2.30] | 1.65 [0.82, 3.29] | 0.64 [0.33, 1.23] | 0.56 [0.28, 1.13] | 0.65 [0.26, 1.59] |
| Professionals | 0.63 [0.34, 1.16] | 0.87 [0.52, 1.46] | 1.36 [0.79, 2.33] | 1.76† [0.92, 3.37] | 0.59† [0.32, 1.07] | 0.82 [0.45, 1.49] | 1.28 [0.60, 2.72] |
| Community and Personal Service Workers | 1.13 [0.57, 2.25] | 0.90 [0.48, 1.66] | 1.46 [0.79, 2.70] | **2.09*** [1.02, 4.31] | 0.75 [0.37, 1.55] | 1.30 [0.67, 2.53] | 1.09 [0.40, 2.99] |
| Clerical and Administrative Workers | **0.36*** [0.16, 0.83] | 0.72 [0.39, 1.33] | 0.71 [0.37, 1.38] | 1.65 [0.80, 3.42] | 0.69 [0.34, 1.39] | 0.59 [0.29, 1.21] | 1.70 [0.74, 3.88] |
| Sales Workers | 0.52 [0.20, 1.38] | 1.74 [0.83, 3.62] | 0.50 [0.21, 1.18] | 1.69 [0.69, 4.10] | 0.91 [0.36, 2.30] | 1.09 [0.47, 2.49] | 1.42 [0.41, 4.97] |
| Technician and Trade, Machinery Operators and Drivers, Labourers | 1.00 (ref.) | 1.00 (ref.) | 1.00 (ref.) | 1.00 (ref.) | 1.00 (ref.) | 1.00 (ref.) | 1.00 (ref.) |
| ***Industry*** | | | | | | | |
| Health Care and Social Assistance | 0.87 [0.48, 1.59] | 1.42 [0.92, 2.20] | 1.35 [0.87, 2.11] | 0.90 [0.53, 1.51] | 1.27 [0.76, 2.12] | 0.70 [0.40, 1.22] | 1.12 [0.60, 2.11] |
| Education and Training | 1.57 [0.85, 2.92] | 0.87 [0.51, 1.49] | 1.03 [0.61, 1.75] | 1.04 [0.58, 1.86] | 1.02 [0.56, 1.86] | 0.73 [0.38, 1.39] | 1.84† [0.97, 3.48] |
| Retail Trade | 1.55 [0.67, 3.58] | 1.39 [0.71, 2.73] | 1.69 [0.84, 3.40] | 1.36 [0.63, 2.92] | 0.97 [0.42, 2.24] | 1.09 [0.51, 2.32] | 0.55 [0.18, 1.67] |
| Accommodation and Food Services | 1.07 [0.46, 2.51] | 1.58 [0.78, 3.20] | **2.40*** [1.23, 4.68] | 1.23 [0.58, 2.63] | 1.62 [0.73, 3.56] | 0.58 [0.25, 1.36] | 0.54 [0.16, 1.80] |
| Other Industries | 1.00 (ref.) | 1.00 (ref.) | 1.00 (ref.) | 1.00 (ref.) | 1.00 (ref.) | 1.00 (ref.) | 1.00 (ref.) |
| ***Employment type*** | | | | | | | |
| Casual | 0.86 [0.51, 1.45] | **0.57*** [0.36, 0.89] | 0.95 [0.62, 1.47] | 1.60† [0.99, 2.56] | 0.91 [0.55, 1.50] | 1.13 [0.69, 1.85] | **0.40**** [0.21, 0.77] |
| Part-time | 0.73 [0.41, 1.29] | 0.77 [0.50, 1.19] | 0.71 [0.45, 1.10] | 0.78 [0.47, 1.30] | 0.71 [0.43, 1.18] | 1.14 [0.68, 1.91] | **0.30**** [0.15, 0.57] |
| Other (e.g. partner, contractor) | 1.06 [0.22, 5.17] | 1.84 [0.59, 5.77] | 0.66 [0.18, 2.49] | 1.33 [0.33, 5.40] | 1.24 [0.30, 5.07] | 1.31 [0.26, 6.63] | 0.76 [0.14, 4.03] |
| Full-time | 1.00 (ref.) | 1.00 (ref.) | 1.00 (ref.) | 1.00 (ref.) | 1.00 (ref.) | 1.00 (ref.) | 1.00 (ref.) |
| ***Current work status*** | | | | | | | |
| Not Working | 0.73 [0.36, 1.51] | 0.84 [0.47, 1.49] | **1.92*** [1.09, 3.36] | **2.01*** [1.12, 3.58] | 0.89 [0.47, 1.68] | 0.82 [0.42, 1.58] | **0.29**** [0.13, 0.63] |
| Usual workplace | 0.64† [0.38, 1.08] | 0.76 [0.50, 1.16] | 0.87 [0.56, 1.35] | **0.52**** [0.32, 0.85] | **0.49**** [0.30, 0.80] | **0.50*** [0.30, 0.83] | **0.11**** [0.06, 0.20] |

| | | | | | | | |
|---|---|---|---|---|---|---|---|
| Both working from home and usual workplace | 0.49† [0.22, 1.11] | 0.71 [0.39, 1.29] | 0.64 [0.34, 1.21] | 0.70 [0.36, 1.39] | 0.83 [0.44, 1.56] | 0.54 [0.26, 1.13] | 0.89 [0.49, 1.61] |
| Working from home | 1.00 (ref.) | 1.00 (ref.) | 1.00 (ref.) | 1.00 (ref.) | 1.00 (ref.) | 1.00 (ref.) | 1.00 (ref.) |
| **Health** | | | | | | | |
| ***Pre-existing medical conditions*** | | | | | | | |
| None | 1.00 (ref.) | 1.00 (ref.) | 1.00 (ref.) | 1.00 (ref.) | 1.00 (ref.) | 1.00 (ref.) | 1.00 (ref.) |
| One | 1.25 [0.76, 2.06] | 1.12 [0.75, 1.67] | 0.96 [0.63. 1.48] | **0.58*** [0.35, 0.95] | 1.31 [0.81, 2.10] | 0.82 [0.50, 1.34] | 0.96 [0.54, 1.68] |
| Two or more | 1.06 [0.55, 2.04] | 0.69 [0.39, 1.22] | 1.23 [0.71, 2.13] | **0.48*** [0.26, 0.89] | 1.43 [0.78, 2.64] | 0.95 [0.51, 1.77] | 1.04 [0.50, 2.16] |
| ***Psychological distress*** | | | | | | | |
| Low to None | 1.00 (ref.) | 1.00 (ref.) | 1.00 (ref.) | 1.00 (ref.) | 1.00 (ref.) | 1.00 (ref.) | 1.00 (ref.) |
| High to Moderate | 1.03 [0.65, 1.63] | **2.09**** [1.47. 2.98] | **2.65**** [1.85, 3.80] | **2.31**** [1.55, 3.46] | 1.43† [0.94, 1.25] | **1.58*** [1.03, 2.40] | **1.63*** [1.01, 2.65] |
| ***Self-rated health*** | | | | | | | |
| Poor to Good | 0.96 [0.64, 1.45] | 1.12 [0.81, 1.55] | 0.92 [0.66, 1.28] | **1.54*** [1.06, 2.25] | 1.22 [0.84, 1.78] | 1.41† [0.96, 2.08] | 0.98 [0.62, 1.54] |
| Very Good to Excellent | 1.00 (ref.) | 1.00 (ref.) | 1.00 (ref.) | 1.00 (ref.) | 1.00 (ref.) | 1.00 (ref.) | 1.00 (ref.) |
| **Finance** | | | | | | | |
| ***Financial stress*** | | | | | | | |
| Yes | 1.44 [0.89, 2.32] | 1.15 [0.78, 1.70] | 1.37 [0.92, 2.02] | **1.77**** [1.16, 2.70] | **2.13**** [1.38, 3.29] | 1.40 [0.90, 2.18] | **1.71*** [1.01, 2.88] |
| No | 1.00 (ref.) | 1.00 (ref.) | 1.00 (ref.) | 1.00 (ref.) | 1.00 (ref.) | 1.00 (ref.) | 1.00 (ref.) |
| ***Financial resources*** | | | | | | | |
| No | 1.54 [0.85, 2.80] | 1.19 [0.71, 2.00] | 1.26 [0.75, 2.11] | 1.39 [0.80, 2.41] | 1.16 [0.64, 2.08] | 0.99 [0.56, 1.75] | **2.49*** [1.20, 5.16] |
| Yes | 1.00 (ref.) | 1.00 (ref.) | 1.00 (ref.) | 1.00 (ref.) | 1.00 (ref.) | 1.00 (ref.) | 1.00 (ref.) |
| **Social interactions** | | | | | | | |
| ***Occasions spending time with others in-person*** | | | | | | | |
| None | 1.16 [0.75, 1.80] | 1.20 [0.84, 1.71] | 1.19 [0.83, 1.71] | 1.08 [0.72, 1.62] | 0.78 [0.51, 1.19] | 1.16 [0.75, 1.78] | 1.29 [0.80, 2.08] |
| At least once | 1.00 (ref.) | 1.00 (ref.) | 1.00 (ref.) | 1.00 (ref.) | 1.00 (ref.) | 1.00 (ref.) | 1.00 (ref.) |
| ***Meetings attended*** | | | | | | | |
| None | 0.89 [0.57, 1.39] | 1.03 [0.72, 1.45] | 1.07 [0.75, 1.53] | 0.95 [0.63, 1.44] | 1.13 [0.75, 1.70] | 1.04 [0.68, 1.58] | 0.83 [0.51, 1.36] |
| At least one | 1.00 (ref.) | 1.00 (ref.) | 1.00 (ref.) | 1.00 (ref.) | 1.00 (ref.) | 1.00 (ref.) | 1.00 (ref.) |
| ***Made or received telephone / online calls*** | | | | | | | |
| Fewer than seven occasions | 1.09 [0.73, 1.61] | 1.13 [0.83, 1.54] | 0.82 [0.59, 1.14] | 1.13 [0.78, 1.63] | 0.99 [0.68, 1.43] | 0.80 [0.55, 1.17] | 1.01 [0.66, 1.56] |

| | | | | | | | |
|---|---|---|---|---|---|---|---|
| Seven or more occasions | 1.00 (ref.) | 1.00 (ref.) | 1.00 (ref.) | 1.00 (ref.) | 1.00 (ref.) | 1.00 (ref.) | 1.00 (ref.) |
| **Survey mode** | | | | | | | |
| Online | 1.02 [0.59, 1.79] | **0.42**** [0.26, 0.67] | **0.41**** [0.25, 0.66] | 1.20 [0.73, 1.99] | **0.55*** [0.32, 0.94] | 0.96 [0.57, 1.61] | 1.01 [0.54, 1.90] |
| Telephone | 1.00 (ref.) | 1.00 (ref.) | 1.00 (ref.) | 1.00 (ref.) | 1.00 (ref.) | 1.00 (ref.) | 1.00 (ref.) |

†0.05≤p<0.1, *0.01≤p<0.05, **p<0.01.
